# An Aberration Detection-Based Approach for Sentinel Syndromic Surveillance of COVID-19 and Other Novel Influenza-Like Illnesses

**DOI:** 10.1101/2020.06.08.20124990

**Authors:** Andrew Wen, Liwei Wang, Huan He, Sijia Liu, Sunyang Fu, Sunghwan Sohn, Jacob A. Kugel, Vinod C. Kaggal, Ming Huang, Yanshan Wang, Feichen Shen, Jungwei Fan, Hongfang Liu

## Abstract

Coronavirus Disease 2019 (COVID-19) has emerged as a significant global concern, triggering harsh public health restrictions in a successful bid to curb its exponential growth. As discussion shifts towards relaxation of these restrictions, there is significant concern of second-wave resurgence. The key to managing these outbreaks is early detection and intervention, and yet there is significant lag time associated with usage of laboratory confirmed cases for surveillance purposes. To address this, syndromic surveillance can be considered to provide a timelier alternative for first-line screening. Existing syndromic surveillance solutions are however typically focused around a known disease and have limited capability to distinguish between outbreaks of individual diseases sharing similar syndromes. This poses a challenge for surveillance of COVID-19 as its active periods are tend to overlap temporally with other influenza-like illnesses. In this study we explore performing sentinel syndromic surveillance for COVID-19 and other influenza-like illnesses using a deep learning-based approach. Our methods are based on aberration detection utilizing autoencoders that leverages symptom prevalence distributions to distinguish outbreaks of two ongoing diseases that share similar syndromes, even if they occur concurrently. We first demonstrate that this approach works for detection of outbreaks of influenza, which has known temporal boundaries. We then demonstrate that the autoencoder can be trained to not alert on known and well-managed influenza-like illnesses such as the common cold and influenza. Finally, we applied our approach to 2019-2020 data in the context of a COVID-19 syndromic surveillance task to demonstrate how implementation of such a system could have provided early warning of an outbreak of a novel influenza-like illness that did not match the symptom prevalence profile of influenza and other known influenza-like illnesses.

## Introduction

### Mitigating COVID-19 Resurgence Risk via Syndromic Surveillance

The fast spread of coronavirus disease 2019 (COVID-19), caused by severe acute respiratory syndrome coronavirus 2 (SARS CoV-2), has resulted in a worldwide pandemic with high morbidity and mortality rates^1-3^. To limit the spread of the disease, various public health restrictions have been deployed to great effect, but as of May 2020, international discussion has begun shifting towards relaxation of these restrictions. A key concern is, however, any subsequent resurgence of the disease^4-6^, particularly given that the disease has already become endemic within localized regions of the world^7^. This issue further exacerbated by significant undertesting, where estimates have found that more than 65% of infections were undocumented^8,9^. Additionally, increasing levels of resistance and non-adherence to these restrictions has greatly increased resurgence risk.

A key motivation behind the initial implementation of public health restrictions was to sufficiently curb the case growth rate so as to prevent overwhelming hospital capacities^10,11^. While the situation has been substantially improved, a resurgent outbreak will present much the same threat^11^. Indeed, second-wave resurgence has already been observed in Hokkaido Japan after public health restrictions were relaxed, and these restrictions were re-imposed a mere month after being lifted^12^. Additionally, from a healthcare provider perspective, significant nosocomial transmission rates for the disease have been found despite precautions^13-15^, a significant concern as many of the risk factors in terms of severity and mortality for COVID-19^2,16^ can be commonly found within an in-hospital population. To avoid placing an even greater burden on already strained hospital resources, it is important that healthcare institutions respond promptly to any outbreaks and modify admission criteria for non-emergency cases appropriately. For both reasons, it is critical to detect outbreaks as early as possible so as to contain them prior to requiring reinstitution of these extensive public health restrictions.

Early detection is, however, no mean feat. Reliance on laboratory confirmed COVID-19 cases to perform surveillance introduces significant lag time after the beginning of the potential shedding period as symptoms must first present themselves^17,18^ and be sufficiently severe to warrant further investigation, before test results are received. This is further complicated by limited test reliability, with RT-PCR tests having an estimated sensitivity of 71%^19^, and serological tests, despite having high reported specificity, having significant false positive rates due to the relatively low prevalence of COVID-19 amongst the population. Moreover, asymptomatic carriers, which in some studies have been found to reach as much as 50-75% of the actual case population^20-22^, present significant risk, particularly amongst the healthcare provider population.

It is therefore evident that any surveillance solution relying purely on laboratory-confirmed cases will suffer from a significant temporal delay as compared to when the transmission event actually occurs, suggesting that a syndromic surveillance solution may be necessary^23^. In this study, we aim to perform computational syndromic surveillance for novel influenza-like illnesses such as COVID-19 amongst a hospital’s patient population (comprising both inpatient and outpatient settings) to detect outbreaks and prompt investigation in advance of actual confirmation of cases.

### Syndromic Surveillance for COVID-19 and Other Novel Influenza-Like Illnesses

Digital syndromic aberration surveillance systems came to the forefront of national scientific attention for bioterrorism preparedness purposes^24^, particularly in the wake of the anthrax attacks in the fall of 2001^25^. Such systems, however, were quickly noted to be also of use in clinical and public health settings^26^. Approaches that have been explored for this task^27^ include usage of simple statistical thresholds on raw frequency or prevalence data, to statistical modeling and visualization approaches such as Cumulative Sums (CUSUM), Exponentially Weighted Moving Averages (EWMA), and autoregressive modeling^28-33^. More specifically to syndromic surveillance of influenza-like illnesses (ILI), at a national level, the United States Centers for Disease Control and Prevention operates the ILInet, a national statistical syndromic surveillance solution deriving its data from reports of fever, cough, and/or sore throat without a known non-influenza cause within outpatient settings^28^.

While generally effective, many of these approaches are limited in granularity to a syndrome level: that is to say they perform surveillance of the frequencies or prevalence of a particular syndrome as a whole, but do not make a distinction amongst individual diseases that share similar syndromes. This is an issue for our task at hand as COVID-19’s syndrome very closely resembles that of many other seasonal diseases such as influenza, the common cold, or even allergic reactions. As such, while an outbreak of a novel influenza-like illness like COVID-19 may be registered in these surveillance systems, they may be difficult to discern if the outbreak temporally overlaps with known seasonal illnesses sharing the same syndrome (e.g. if they begin at the height of the influenza season), and the ongoing outbreak may be misattributed to the more benign seasonal disease.

The underlying symptom prevalence amongst positive cases of influenza-like illnesses is, however, perceptibly different. For instance, while the symptom prevalence distribution for positive cases of influenza amongst the hospitalized, vaccinated, sub-50, population is 98%, 88%, 83%, 87%, and 96% for cough, fever, headache, myalgia, and fatigue respectively^34^, the distribution for the same symptoms is 59%, 99%, 7%, 35%, and 70% respectively for hospitalized COVID-19 positive cases^13^. As an outbreak of COVID-19 will likely affect the background symptom prevalence distribution in a different manner than an outbreak of influenza, we theorize that an approach incorporating symptom prevalence distributions as part of its input data as opposed to the frequency/prevalence of the syndrome as a whole will be able to perform this differentiation and as such suppress outbreaks of known, relatively benign, seasonal diseases at the user’s discretion.

Machine learning approaches can be used to perform this anomaly detection task. In this study, we adapted one such commonly used approach within the general domain, autoencoders^35-37^, for our syndromic surveillance task. An autoencoder (also commonly termed a “Replicator Neural Network”) is a neural network trained in a self-supervised manner to first encode the input into a lower-dimensional form, and then decode this lower-dimension form to reconstruct the input^38^. In other words, a trained autoencoder learns two functions, an encoding function and a decoding function, such that given an input *x, encode*(*x*) = *y, decode*(*y*) ≅ *x*, |*x*| > |*y*| and *x* ≠ *y*. A natural property of autoencoders is that their encoding and decoding functions only function properly for input data that is similar to the data for which it is trained: data that differs in its input features will fail to be successfully reconstructed such that *decode*(*y*) ≠ *x*.

For the purposes of syndromic surveillance, we theorize that the autoencoder approach can be adapted: given a distribution of syndromic prevalence within the clinic, we would expect that distribution to change significantly should an outbreak occur. This implies that during an actual outbreak, the reconstruction error would increase perceptibly as compared to during normal time periods and can thus be plotted against time to provide a readily interpretable visualization of an outbreak of a novel influenza-like illness.

In other words, to accomplish the COVID-19 and other novel influenza-like illness syndromic surveillance task, we propose that:

1. By mining the raw mentions of symptoms within a syndrome of interest through a NLP-based approach, we can estimate the prevalence of individual symptoms amongst the overall patient population in a timely manner
2. By delineating certain time periods as “normal” (i.e. no outbreaks of surveilled target of interest) for autoencoder training purposes, the resulting model can be used to perform syndromic surveillance by measuring the error score of any given day’s input symptom prevalence distribution. Crucially to the COVID-19 and novel ILI detection task itself, “normal” time periods can also contain outbreaks of seasonal influenza, which should lead the model to learn the appropriate symptom prevalence distributions so as to not have elevated errors during typical influenza seasons.

In this study, we explore this approach for computational syndromic surveillance with the goal of enabling early detection of outbreaks of COVID-19 and other novel influenza-like illnesses, particularly during periods of heightened seasonal influenza-like-illness activity.

## Results

### Overview

The true beginnings of the COVID-19 pandemic within the United States is still a subject of much contention, with the date being pushed earlier as investigation continues^39^. As such, it is difficult to directly validate any conclusions about the viability of autoencoder-based syndromic surveillance for COVID-19. As such, we validated our approach incrementally through a three-phase approach:

1. Validating the utility and accuracy of autoencoder-based anomaly detection for syndromic surveillance on a disease with known outbreak time periods
2. Validating that given appropriate training data, our autoencoder model can effectively learn the symptom distributions of outbreaks of COVID-19’s common seasonal differentials such as influenza, allergies and the common cold within its underlying model, i.e. that it is capable of suppressing outbreaks of these other, known, seasonal illnesses from its resulting signal
3. Applying an autoencoder based anomaly detection approach to syndromic surveillance of COVID-19 over the past year of data and evaluating the resulting error plot against currently known key dates for the COVID-19 pandemic

### Autoencoder-Based Anomaly detection is Viable for Syndromic Surveillance Tasks

To validate the utility and accuracy of autoencoder-based anomaly detection for syndromic surveillance, we chose syndromic surveillance of influenza seasons as the target task. This task was chosen primarily due to two factors: 1) its relatively well-defined outbreak periods (available both at a national and state level via the CDC Morbidity and Mortality Weekly Reports^40-47^ and the CDC Influenza-Like-Illness (ILI) Activity Tracker^48^ respectively) and 2) its similarity in potential input features (due to similar symptom presentations) to our end-goal of performing COVID-19 syndromic surveillance.

In Figure 1, we present the error plot relative to the anomaly threshold of a stacked autoencoder trained using influenza off-season data for the purposes of syndromic surveillance of influenza. We additionally highlight official CDC flu seasons (national level) ^40-47^ in orange, and time periods with heightened (moderate or greater) ILI activity^48^ within the state of Minnesota (from where our data originates) in red.

**Figure 1.**
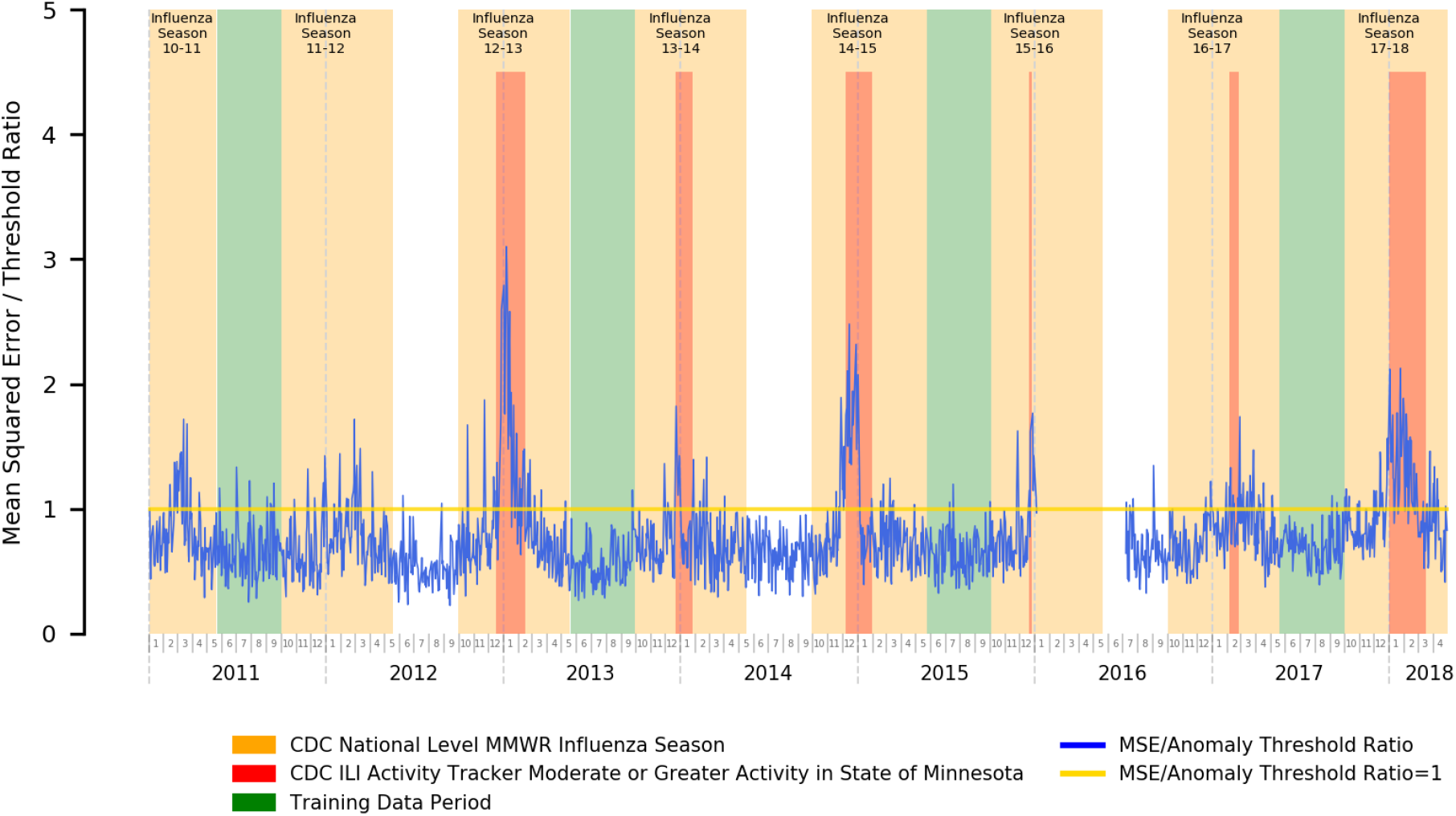
Mean Squared Error Relative to Anomaly Threshold for an Autoencoder Trained for the Influenza Season Detection Task

Our error plots and the close congruence between periods of heightened autoencoder reconstruction error and influenza activity does suggest that our approach is fairly successful at performing the influenza syndromic surveillance task. Of particular note, the magnitude of the reconstruction error is also closely tied to the associated severity of the outbreak, as can be seen in the location of our error peaks relative to state-level ILI activity tracking.

As such, our results here suggest that an autoencoder-based anomaly detection approach to syndromic surveillance is capable of picking up and alerting on the underlying changes in the prevalence of influenza-related symptoms in the practice during influenza season as opposed to the off-season, both in terms of identifying that the underlying distribution of symptom prevalence changed and in reflecting the magnitude of the differences in underlying distribution of symptom prevalence compared to normal time periods within its reconstruction error.

These results are promising for our eventual experiment for COVID-19 syndromic surveillance as the underlying assumptions are similar: COVID-19 and influenza share very similar symptoms, but the underlying distribution of the prevalence of individual symptoms within their respective cases will likely differ. It is expected that an autoencoder will be able to pick up on these prevalence distribution differences in a similar manner to the influenza season vs. offseason variation.

### Autoencoders can be Trained to Suppress Alerting on Outbreaks of Illnesses Sharing Similar Syndromes

COVID-19 syndromic surveillance is severely complicated by its similar presentation and overlapping timeframe with a variety of seasonal illnesses, such as the common cold, allergies, and influenza. To verify that an autoencoder-based COVID-19 syndromic surveillance solution will be functional, we must first verify that, if supplied as part of its training data, outbreaks of these seasonal illnesses will not be reflected in its resulting error plots. To that end, we again use influenza as the target for evaluation here, due to its relatively well-defined temporal boundaries.

In Figure 2, we present the mean squared error plot of a stacked autoencoder trained using data covering three influenza seasons and off-seasons, with the aim of verifying that typical influenza seasons can be suppressed from anomalous readings by incorporating their symptom prevalence distributions as part of training data.

**Figure 2.**
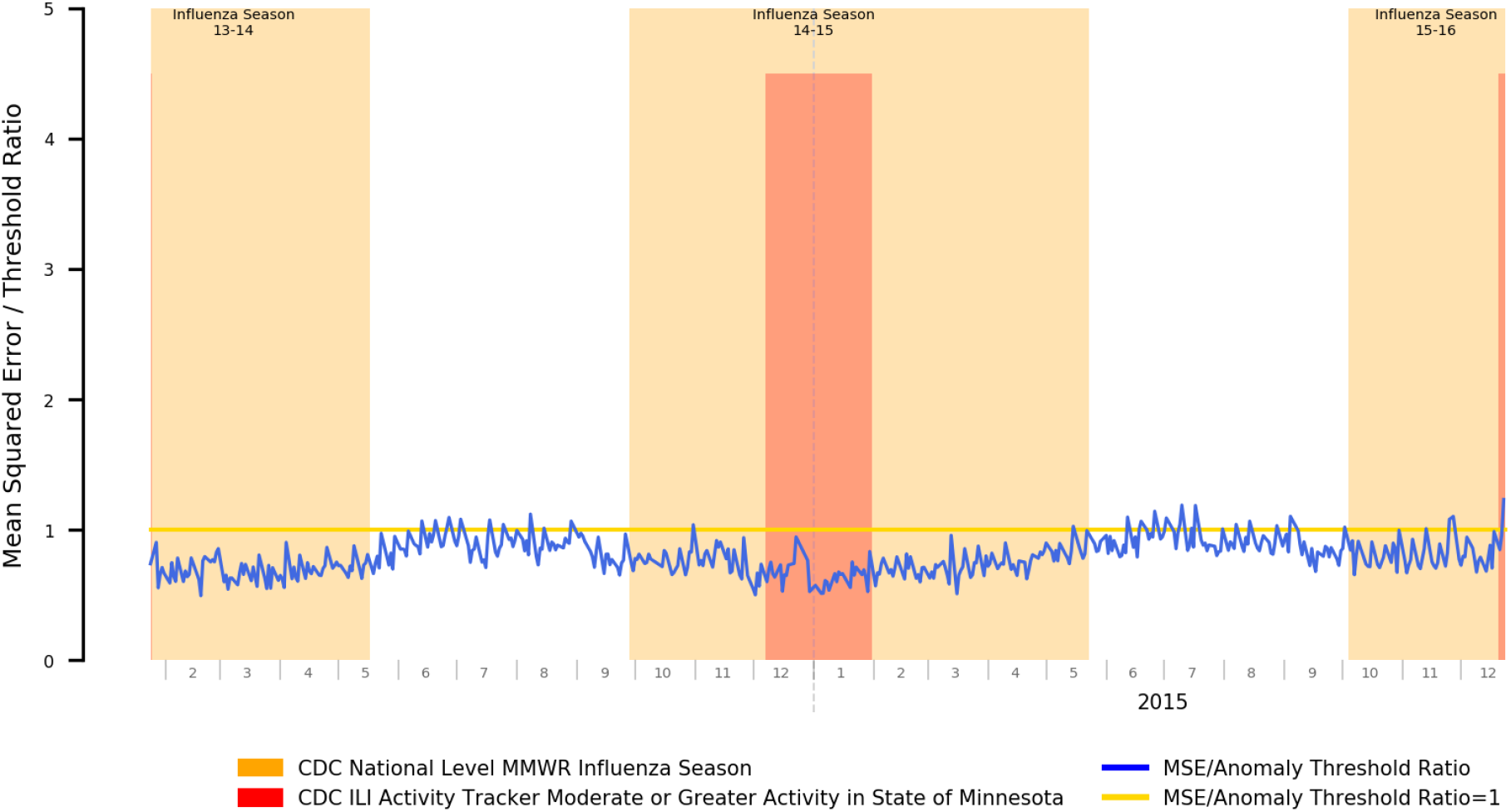
Mean Squared Error Relative to Anomaly Threshold for an Autoencoder Trained on both Influenza Season and Offseason Data

Our results demonstrate that our autoencoder has successfully incorporated symptom prevalence data for influenza and other seasonal diseases with similar differentials occurring within the target period, as can be seen by the relatively consistent reconstruction error throughout the year with peaks being dramatically suppressed in magnitude compared to the highly visible peaks in Figure 1.

### Syndromic Surveillance Viable for Sentinel Detection of Novel Influenza-Like-Illnesses

At this point we have validated that a) an autoencoder reconstruction error-based approach to anomaly detection is capable of reflecting both the occurrence and the magnitude of shifts in underlying symptom prevalence distributions, and b) if included as part of the “normal” training data, autoencoders will successfully reconstruct symptom prevalence distributions occurring during COVID-19’s seasonal differentials. We can thus proceed with the targeted task of this study: syndromic surveillance of the COVID-19 outbreak within the United States, particularly within Olmsted County, Minnesota, the location of the Mayo Clinic Rochester campus.

In Figure 3, we present the mean error plot of a stacked autoencoder trained using a year of both influenza season and off-season data applied to data from June 1^st^, 2019 through April 30^th^, 2020. We additionally annotated the resulting plot with dates pertinent to the COVID-19 epidemic in Minnesota to provide additional context to the detected signals.

**Figure 3.**
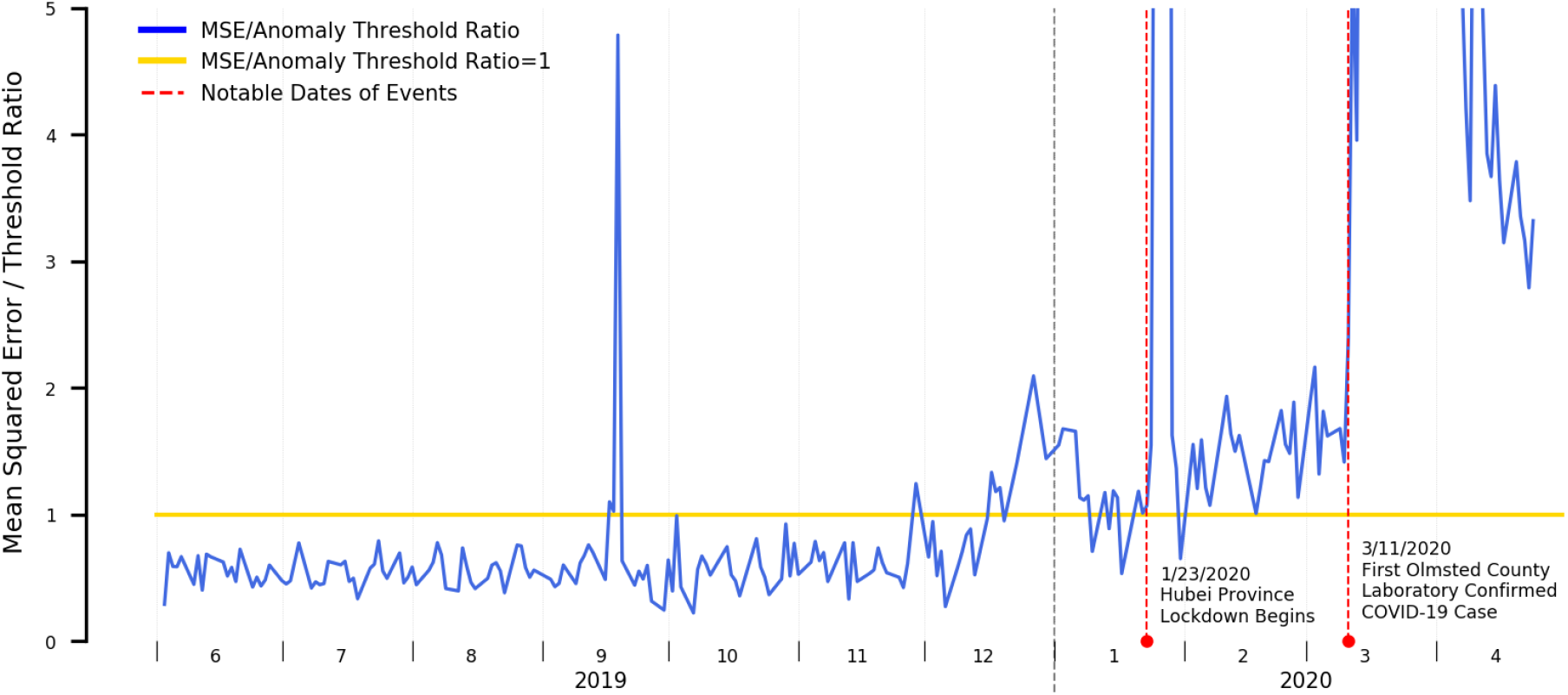
Mean Squared Error Relative to Anomaly Threshold for an Autoencoder Trained on both Influenza Season and Offseason Data

Our results suggest the following with respect to the time period prior to the first laboratory confirmed case in the state of Minnesota:

1. A spike occurring the week of September 15^th^, 2019. We do not believe this is COVID-19 related and will elaborate more on this in the discussion section.
2. A persistent, low level of elevated anomalous signals beginning late December through the first laboratory confirmed COVID-19 case within Olmsted County, Minnesota occurring March 11^th^, 2020. This period is marked by two dramatic spikes occurring January 23^rd^ and March 11^th^ 2020 that we will also discuss in the discussion section. This period of elevated anomalous signals does roughly match the period of heightened state-level ILI activity as reported by the CDC.

When interpreting these results, it is important to note that CDC’s ILI tracker is itself a form of syndromic surveillance and doesn’t explicitly indicate levels of influenza-specific activity, but rather all syndromes with similar symptomatic presentations: specifically, ILInet uses fever, cough, and/or sore throat without a known non-influenza cause as the data through which it performs its tracking^28^. It is therefore expected that our detected anomalous time periods will match, as COVID-19 itself shares many of these symptoms.

The fact that elevated anomalous results appeared in our error plot, however, suggests that the underlying symptom prevalence distributions seen within the clinical practice are atypical of those seen in other influenza seasons: per the second phase of our experiment, we established that “typical” influenza seasons can be suppressed from anomalous readings by incorporating their symptom prevalence distributions as part of training data. We would have thus expected the error rates to have remained largely under the anomaly threshold with no significant peaks, unlike what was observed here.

## Discussion

### Interpreting Anomalous Signals and Potential Attribution Errors

It is important to note with all our results presented here that the anomaly detection component detects anomalies in the input data, i.e. anomalies in the incoming symptom prevalence distributions. Such anomalies can, however, be caused by a variety of external factors and are not necessarily indicative of an outbreak. As such, while such a system can serve as an early-warning system to alert that an anomaly exists as well as the magnitude of such an anomaly, further human investigation is needed to identify the underlying reasons as well as to confirm whether an outbreak is occurring. With reference to our results derived from Figure 3 suggesting a sustained elevated anomalous error rate starting the final week of December through the first laboratory confirmed COVID-19 case, it would therefore be premature to directly conclude that the anomalous time period is attributable to only COVID-19, such a conclusion would only be possible to achieve had laboratory tests been done during that time period. Instead, it serves only as an indicator of the need for additional investigation.

An example of the potential for attribution error can be shown where, in Figure 4, we note that while the periods of elevated error rates for the 2017-2018 influenza season do roughly correspond to the official CDC-determined flu season and periods of heightened ILI activity, starting May of 2018, the error rate rises outside the display range of the chart. This anomaly does, in fact, exist in reality, but is not tied to a renewed outbreak of influenza-like illness. Rather, the Mayo Clinic Rochester clinic migrated EHR systems from its historical GE Centricity-based EHR to the Epic EHR, and the go-live date for clinical operations was May 1^st^. Due to the changes in clinical workflows and associated documentation practices, the underlying distribution of positive symptom prevalence mentions within clinical documentation also dramatically changed, and that anomalous change was appropriately detected.

**Figure 4.**
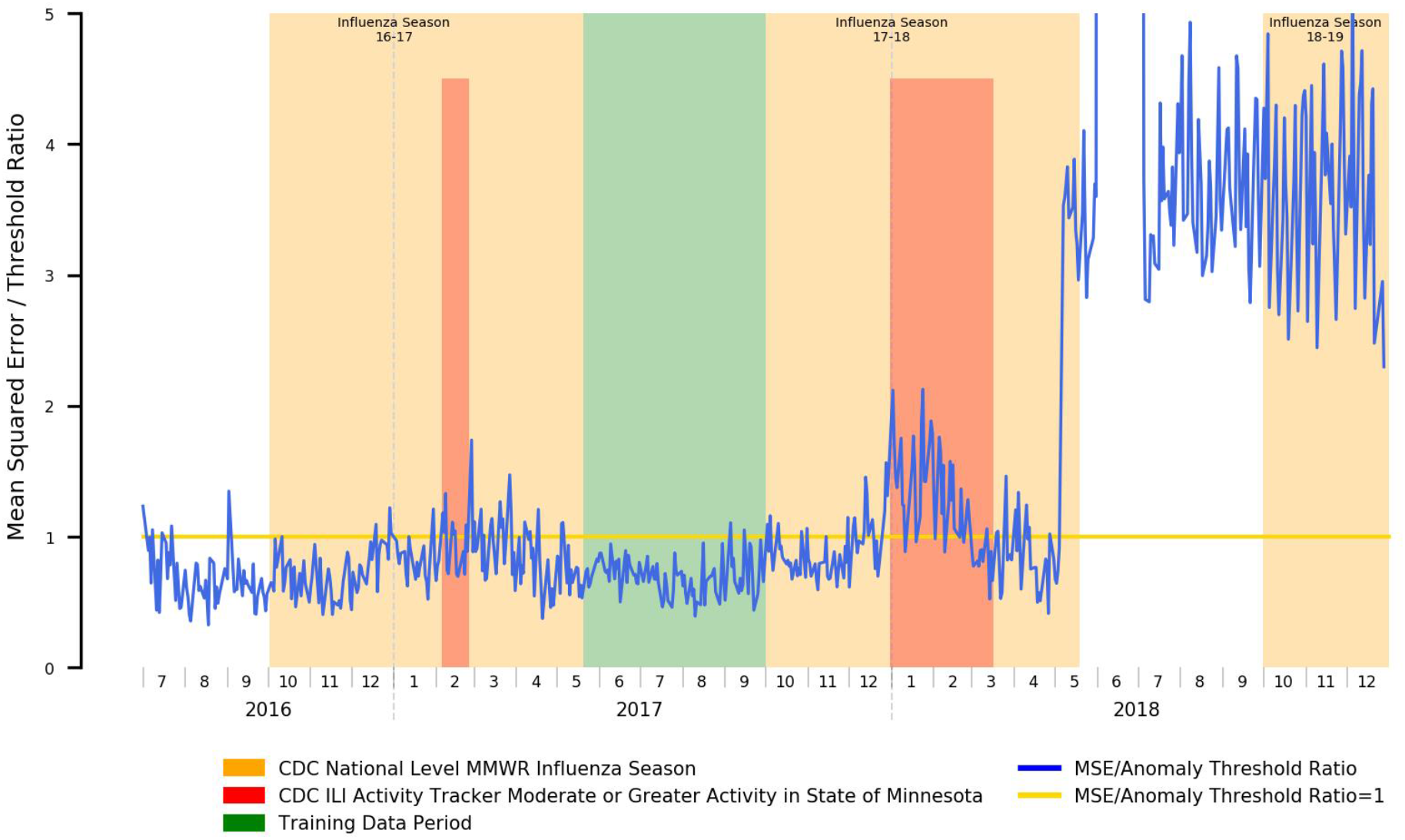
Mean Squared Error Relative to Anomaly Threshold for an Autoencoder Trained for Influenza Season Detection Spanning an EHR Migration Occurring May 2018

A similar phenomenon is reflected in Figure 3. A brief spike in the plotted errors occurs mid-September 2019: further investigation leads us to hypothesize that rather than an outbreak of influenza-like illness during this timeframe, this spike was related to media coverage and associated greater patient concern to a local outbreak of E.coli during this same time period originating from a popularly attended state fair^49^. Similarly, two events that triggered greatly increased media coverage and associated public awareness are highlighted in red, the initial lockdown of the city of Wuhan and Hubei province on January 23^rd^ 2020, the event that originally brought the coronavirus outbreak to the public’s attention, and the first laboratory-confirmed COVID-19 case within Olmsted County, Minnesota on March 11^th^ 2020. Instead of directly attributing the spike to only actual [undiagnosed] COVID-19 cases, the news coverage and increased patient concern likely caused a dramatic increase in patient healthcare engagement, and that increase is likely reflected here with the dramatic spikes. Nevertheless, these “public awareness and concern” spikes are typically obvious, as the spike is sudden, relatively large in magnitude, and are temporally co-located with publicly available news sources.

### COVID-19 Syndromic Surveillance: Retrospective and Prospective Opportunities

Had a syndromic surveillance solution similar to what we established in phase 3 of our experiment existed at the time of the Hubei lockdown, anomalous readings would have appeared far in advance of the actual first laboratory-confirmed case even within the United States, and alert on a possible outbreak a novel influenza-like-illness that did not share similar symptom prevalence distributions as priorly encountered influenza seasons. This information could have been used as an actionable signal for further investigation suggesting a possible spread of COVID-19 within the served community and been a prompt for far more aggressive testing than what was done in practice. From a public health perspective, this could have allowed for earlier intervention and potentially dramatically reduced outbreak magnitude.

From a prospective perspective, such a syndromic surveillance approach can potentially be utilized to provide early warning of future outbreaks, particularly with respect to differentiation from outbreaks of other influenza-like illnesses. As public health restrictions are eased, such capabilities are increasingly critical for detection and early intervention in the case of second-wave outbreaks within the individual hospital’s served communities. It is important to note, however, that clinical workflows with respect to patients presenting with influenza-like illnesses, and by extension documentation practices will have substantially changed in the post COVID-19 era; these changes will be reflected in elevated error rates. Such a discrepancy may be addressed through the application of transfer learning: with a pretrained model similar to that which would be produced from phase 3 of our experiment, limited retraining of the existing model on a month of “normal” data after resumption of full clinical operations might be sufficient to adapt it to the post COVID-19 data distributions.

### Data and Study Limitations

Our study faced several challenges from a data perspective. Firstly, it must be noted that patient profiles significantly change between normal work-week operations and weekends/holidays, which are far more likely to be acute/emergency care. As such, to prevent these from becoming a confounding factor and unduly influencing our anomaly detection error plots, data points relating to weekends, US federal holidays, Christmas Eve and New Year’s Eve were excluded from our datasets. We do not believe that this has affected the validity of our results, further evidenced by the plot in Figure 4, showing that the period of elevated ILI activity that occurred from January through mid-March of 2018 was correctly reflected, while December of 2017 did not display anomalous results, indicating that our model is not simply picking up on proximity to holidays. We will, however, work on incorporating weekend and holiday data as part of our models as part of future work.

Additionally, several limitations within our data sources hampered our efforts to evaluate our methods: as previously noted, anomalies may also be caused by problems with the input data unrelated to the syndromic surveillance task. Specifically, in our case, we faced two major EHR/data platform shifts within our source data that led to irregular disruption of clinical documentation within our data warehouse, one occurring throughout the entirety of Q1 2016, and the other occurring beginning May 1^st^ 2018 and lasting through the first week of July 2018 resulting from Mayo Clinic Rochester’s migration to the Epic EHR. The training datasets and results presented thus excluded these time periods (except for illustrative purposes in Figure 4) as they are known to be anomalous with the reasons for the anomaly being irrelevant to our target tasks (e.g. reasons for anomaly include changes in documentation practices affecting NLP-based prevalence, metadata changes, etc.)

Finally, the fact that an EHR migration did occur significantly hampers the amount of pre-COVID-19 data available for training purposes for phase 3 of our experiment. Due to documentation practice shifts we must use Epic data as part of our training data, and due to the data source disruption as a result of this migration, we were limited to data beginning August of 2018. As part of future work, we thus aim to further validate our model on other sites within the Mayo Clinic enterprise that switched EHR systems in 2016, so as to have a greater amount of training data.

From a methodological perspective, we were constrained in available methodological choices by the need for methods to be unsupervised and/or self-supervised (using “normal” data): given our task to detect novel influenza-like-illnesses of unknown symptom prevalence distributions, it is not feasible to procure labeled “anomalous” data for supervised learning approaches. It is nevertheless important to note that the autoencoder approach is only one of many existing approaches that have been utilized for anomaly detection within the general domain. Other approaches commonly used in this space include k-means clustering^50-52^, one-class SVMs^52-54^, Bayesian networks^55^, as well as more traditional statistical approaches such as the chi-square test^56^ and principal component analysis^57^. In many systems, such approaches are not taken in isolation, but are rather used in conjunction with others to perform specific sub-components of the anomaly detection task or to provide multiple features for downstream analysis^51,53,58,59^. Our study is not intended to perform a comprehensive benchmarking of available methods, and we have not included comparative metrics here given that we have achieved workable results with only an autoencoder approach. Nevertheless, it may be worth exploring usage and/or integration of many of these other models to improve discriminative power and denoise the signal, and we have left such exploration to future work.

## Methods

We present an overview of our experimental procedure in Figure 5, and outline each step in detail within the ensuing subsections.

**Figure 5.**
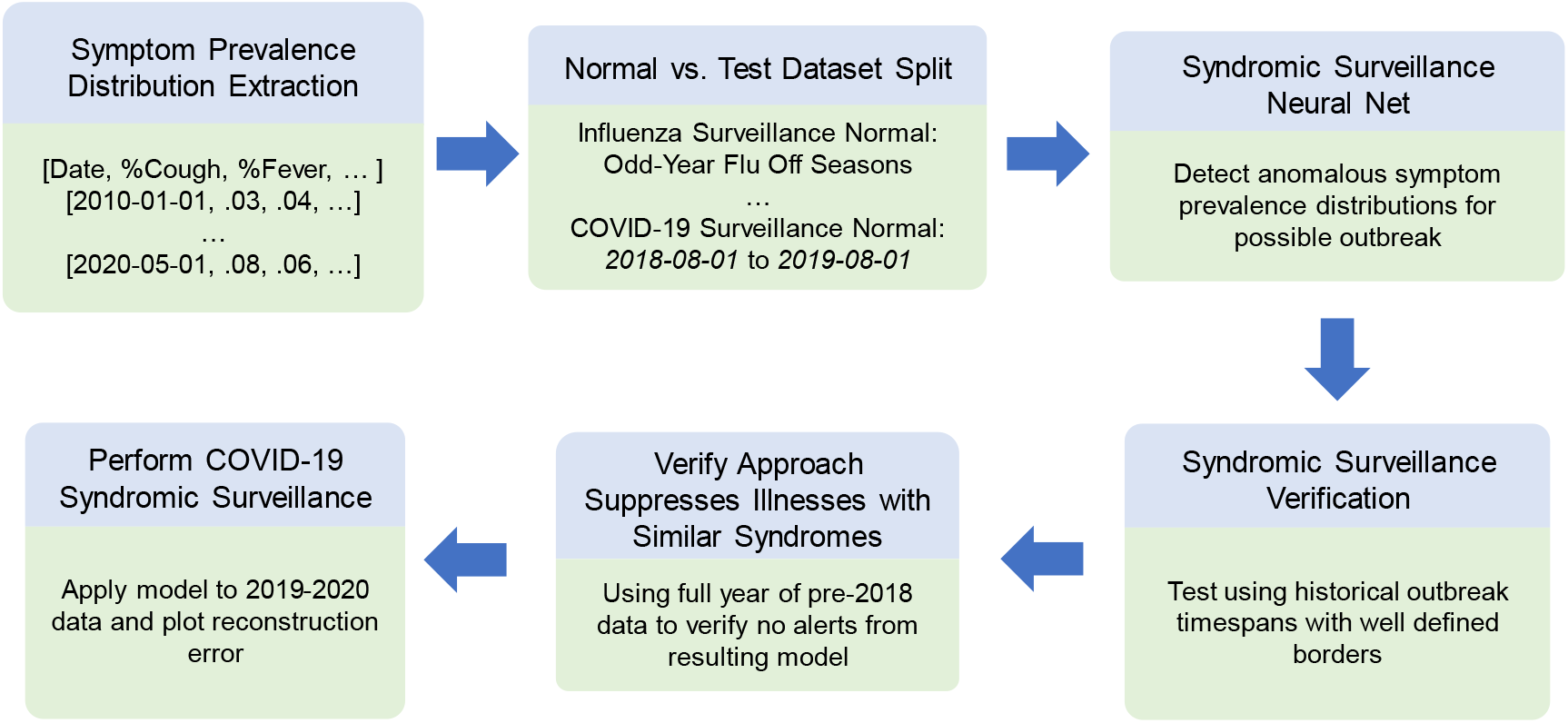
Experimental Procedure Overview

### Sign/Symptom Extraction

Sign and symptom extraction via natural language processing was accomplished via the MedTagger NLP engine^60,61^. The signs and symptoms chosen were selected via a literature review conducted in early March 2020 for known COVID-19 and influenza symptoms^13,62^. Specifically, mentions of Abdominal Pain, Appetite Loss, Diarrhea, Dry/Nonproductive Cough, Dyspnea, Elevated LDH, Fatigue, Fever, Ground-Glass Opacity Pulmonary Infiltrates, Headaches, Lymphopenia, Myalgia, Nasal Congestion, Patchy Pulmonary Infiltrates, Prolonged Prothrombin Time, and Sore Throat were used for all three experiments. Additionally, explicit mentions of influenza were used for phase 2 (establishing baseline/incorporating influenza seasons as part of “normal” symptom prevalence distributions) and phase 3 (COVID-19 surveillance task) of our experiment. Only positive present NLP artifacts with the patient as the subject were retained.

### Symptom Prevalence Distribution Dataset

Clinical documentation generated from January 1^st^ 2011 through May 1^st^ 2020 was utilized as part of this study, with the exclusions detailed within the Data Limitations subsection within our Discussion section (January 1^st^ – July 1^st^ 2016, May 1^st^ – July 7^th^ 2018). For each day within this range, a symptom prevalence feature vector was generated, where each item in the vector corresponds to the symptom prevalence of one of the symptoms of interest for that day. We define symptom prevalence on any given day as the number of unique patients that had a clinical document generated that day containing a NLP artifact corresponding to that symptom (that was positive, present, and had the patient as the subject) divided by the number of unique patients that had at least 1 clinical document generated on that day.

This dataset was then subdivided into different training and plotting (for simulated surveillance purposes) definitions for each of the tasks at hand. We have provided a summary of these divisions in Table 1.

**Table 1.**
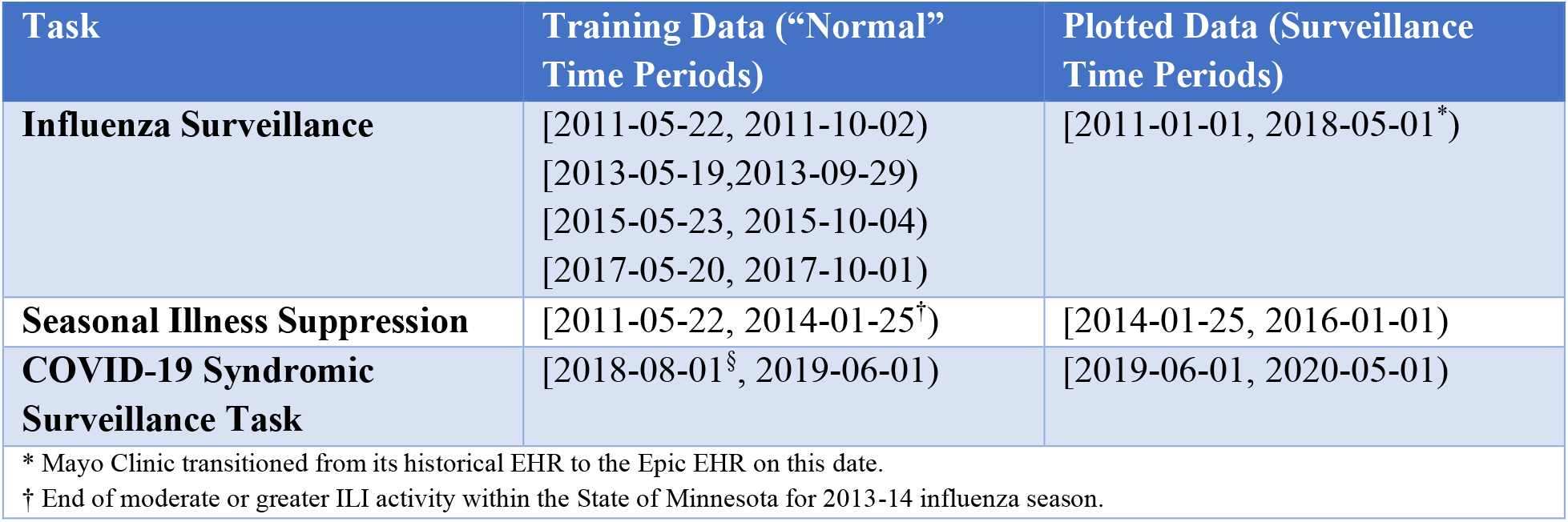
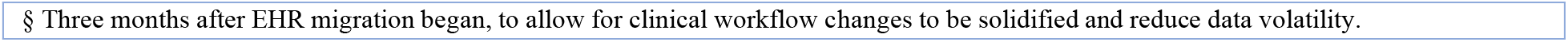
Task-Specific Training and Plotted Data Divisions

### Autoencoder Architecture and Implementation

Our neural network was implemented in Java via the DL4J deep learning framework^63^. For our purposes we used a 5-layer fully-connected stacked autoencoder consisting of [INPUT_DIM, 14, 12, 14, INPUT_DIM] nodes in each respective layer, where INPUT_DIM refers to the dimensionality of the input data. For influenza detection, this was 16 (excluding influenza prevalence), and for all other tasks, this was 17. The activation function used for all layers was the sigmoid activation function, except for the output layer, which used the identity function, with all inputs being rescaled to the [-1, 1] range. The optimization function used, their associated learning rate, and the l2 regularization penalty was selected via five-fold cross-validation, where optimization function was one of AdaDelta^64^, AdaGrad^65^, or traditional stochastic gradient descent^66^ and their respective learning rate was selected from 100 randomly sampled points from the range [0.0001, 0.01], with the exception of AdaDelta, as it is an adaptive learning rate algorithm, and we instead used the recommended default rho and epsilon of 0.95 and 0.000001 respectively. An L2 regularization penalty^67^ was selected from 100 random samples in the range [0.00001, 0.001]. The cost function used was mean squared error. For all model training tasks, training was done using the entire train dataset as one batch, over 1000 epochs utilizing early stopping (5 iterations with score improvement < 0.0001) and selecting the model resulting from the epoch that had the best performance against the test dataset.

### Evaluating Influenza Season Detection Capabilities

For training purposes, we used seasonal date ranges as defined in the US CDC released morbidity and mortality weekly report (MMWR) and selected flu offseason for the odd-numbered years between 2010 and 2018 as our training set^40-47^. Specifically, the date ranges used for training were [2011-05-22, 2011-10-02), [2013-05-19,2013-09-29), [2015-05-23, 2015-10-04), and [2017-05-20, 2017-10-01). For these date ranges, all extracted symptom prevalence information was included for training with the exception of explicit mentions of influenza, as that might provide an unwarranted hint for the task to the underlying trained network.

To evaluate this approaches’ effectiveness for influenza season detection, we ran the trained autoencoder on all years from 2011 through May of 2018 (when the Epic EHR migration occurred), and plotted the error, as determined by the mean-squared error between the supplied input feature set and the network’s outputs, with a particular focus on detected influenza seasons starting on even years.

The best performing model from training was selected, and the anomaly threshold was determined as the mean + 2 standard deviations of the reported errors derived from the test partition resulting from cross-validation of the normal (training) time periods, with errors higher than this value being deemed anomalous.

The errors were plotted and compared against timespans with elevated influenza activity, both at a national level via the official MMWR defined influenza season and in terms of ILI activity for the state of Minnesota as reported by the CDC ILInet. The distinction is important as while the CDC MMWR reports a national level influenza season, the actual periods of elevated activity differ from state to state, and we would only truly be able to detect anomalies when influenza activity is actually elevated within Minnesota, as that is the source for our data.

### Evaluating Autoencoder Capability to Embed Influenza Season Data as “Normal”

In this phase, we use data from May 22^nd^ 2011 (the end date of the 2010-2011 influenza season) through January 25^th^ 2014 (the end date for observed moderate-or-greater ILI activity in the state of Minnesota for the 2013-2014 influenza season) as our training set.

Unlike in the previous phase, the prevalence of influenza mentions is included within the feature set for training to supply explicit knowledge about the occurrence of and the magnitude of ongoing influenza seasons. Additionally, to ensure a balance in examples, we sampled from the influenza off-seasons such that the number of off-season examples corresponded to the number of in-season examples.

Once training using this dataset was completed, we then ran this new autoencoder model on all data between January 25^th^ 2014 and January 1^st^ 2016 and plotted the mean squared error between the supplied input and the autoencoder’s resultant output, with a focus on even years.

The anomaly threshold was again set to the mean + 2 standard deviations of the test partition error during the training time period and the resultant anomalous spans were used to evaluate the autoencoder’s capability to embed influenza and other seasonal differential data.

Applying Autoencoder-Based Anomaly detection for COVID-19 Syndromic Surveillance In this phase, we use data from August of 2018 through June of 2019 (Exclusive) as our “normal” training data. Again, we ensure a 50/50 balance of influenza in-season and off-season examples in our dataset prior to partitioning the data for cross-validation. As with our previous experiments, the anomaly threshold was set to the mean + 2 standard deviations of the test partition error during the training time period.

The resulting model was run on data from June of 2019 through present, and the resulting errors were plotted for further analysis.

## Data Availability

Due to the results of the symptom extraction process being considered protected health information, data is not available as it would be difficult to distribute to anyone not engaged in an IRB-approved collaboration with the Mayo Clinic.
The NLP engine and associated algorithm used to extract ILI symptoms as described in this study is available within the MedTagger project (https://www.github.com/OHNLP/MedTagger). Please consult the Wiki and README file accessible from the linked page for instructions on how to use for the COVID-19 use case.
The aberration detection/sentinel syndromic surveillance component has been decoupled from institutional data sources and is available at https://github.com/OHNLP/AEGIS. As this is an active project undergoing improvement and new features that may lead to changes in the underlying code inconsistent with what was described in this manuscript, we have tagged the codebase as described in this manuscript with the COVID19 tag.

https://github.com/OHNLP/AEGIS

https://github.com/OHNLP/MedTagger

## Acknowledgements

Research reported in this publication was supported by the National Center for Advancing Translational Science of the National Institutes of Health under award number U01TR002062. The content is solely the responsibility of the authors and does not necessarily represent the official views of the National Institutes of Health.

## Code Availability

The NLP engine and associated algorithm used to extract ILI symptoms as described in this study is available within the MedTagger project (https://www.github.com/OHNLP/MedTagger). Please consult the Wiki and README file accessible from the linked page for instructions on how to use for the COVID-19 use case.

The aberration detection/sentinel syndromic surveillance component has been decoupled from institutional data sources and is available at https://github.com/OHNLP/AEGIS. As this is an active project undergoing improvement and new features that may lead to changes in the underlying code inconsistent with what was described in this manuscript, we have tagged the codebase as described in this manuscript with the COVID19 tag.

## Data Availability

Due to the results of the symptom extraction process being considered protected health information, data is not available as it would be difficult to distribute to anyone not engaged in an IRB-approved collaboration with the Mayo Clinic.

## Author Contributions

AW: Designed, implemented study, performed experiments. AW, LW, HH, SL, SF, MH, YW, FS: Determined symptom inclusion/exclusion criteria for NLP algorithm and similar contributions, preparation of NLP algorithm for public distribution, and other miscellaneous project tasks. HH, SL: Generation of graphs and figures as presented in manuscript. AW, SS, JAK, VCK: NLP engine work used for this study, interfacing with institutional data sources. JF, HL: Direction on study design and conceptualization, project leadership. All authors reviewed and contributed expertise to the final manuscript.

